# Covid-19 second dose vaccine uptake and associated factors: A community-based cross-sectional study among adults in Masindi district in Uganda

**DOI:** 10.64898/2026.10.26.26366515

**Authors:** Ronald Kooko, Agnes Alinde, Larissa Ngazet Yangbalet Kezza, Samkeliso G. Shongwe, Rogers Kisame

**Affiliations:** Ministry of Health-Uganda, Department of Integrated Epidemiology, Surveillance and Public Health Emergencies, Kampala, Uganda; Department of Community Health and Behavioral Science, School of public health, Makerere University, Kampala, Uganda; Baylor College of Medicine Children’s Foundation Uganda, Department of Global Health Security, Kampala, Uganda; Reproductive Health Program, Pan African University Life and Earth Sciences Institute (including Health and Agriculture), University of Ibadan, Ibadan, Nigeria

**Keywords:** COVID-19 vaccination, second dose uptake, Masindi district, Uganda

## Abstract

While Uganda has achieved relatively high COVID-19 first dose vaccination rates, completion of the second dose remains low, particularly in Masindi district where only 44.0% of adults who were vaccinated with the first dose proceeded to receive the second dose. This shortfall threatens to undermine the achievement of herd immunity and the effectiveness of the nation’s COVID-19 vaccination campaign. This study aimed to determine the rate of second dose vaccine uptake and assess factors associated with second dose uptake among adults in Masindi district. We conducted a community-based cross-sectional study using multi-stage random sampling approach to select 637 study respondents. Data were collected using a pre-tested interviewer-administered structured questionnaire and analyzed using SPSS v26. Modified Poisson regression was used to identify independent predictors of second dose uptake. Results were reported using adjusted prevalence ratios (APRs) and 95% confidence intervals (CIs), with statistical significance set at p<0.05. Among adults who had received at least one dose of a COVID-19 vaccine, 44.0% had completed the two-dose vaccination schedule. Independent factors significantly associated with higher second dose uptake included high economic status (APR = 2.2, 95% CI: 1.16-3.07), urban residence (APR = 1.3, 95% CI: 1.13-1.62), high perceived risk of COVID-19 (APR = 2.0, 95% CI: 1.59-2.55), chronic illness (APR 1.9; 95% CI: 1.60-2.33), living less than 1 km from a vaccination site (APR = 2.0, 95% CI: 1.49-3.01), Shorter waiting times; under 30 minutes (APR = 1.7, 95% CI: 1.41-2.08), community/religious leaders’ support (APR = 1.7, 95% CI: 1.41-2.16), Community outreach programs (APR = 2.5, 95% CI: 1.99-3.36), and Exposure to vaccination information through radio or TV (APR = 1.5, 95% CI: 1.25–1.88). Vaccine shortages (APR = 0.4, 95% CI: 0.31-0.55), and experiencing of adverse side effects after the first dose (APR = 0.8, 95% CI: 0.71-0.97) were associated with reduced prevalence of receiving a second dose. The findings suggest that COVID-19 vaccine second dose uptake in Masindi district remains low and is associated with interconnected socioeconomic, individual, health system, and community-level factors. Interventions should consider improving access, risk communication, and engagement of community structures are needed to boost vaccine completion rates.

## Introduction

The COVID-19 pandemic, declared a global emergency by the World Health Organization (WHO) in March 2020, has caused unprecedented morbidity, mortality, and socioeconomic disruption worldwide [1]. Vaccination has emerged as the most effective long-term public health intervention to control the pandemic, complementing earlier strategies such as lockdowns, quarantines, and social distancing [2]. Globally, over 13 billion doses have been administered, with approximately 70% of the population receiving at least one dose [3]. However, disparities persist between high-income countries and low- and middle-income countries (LMICs), largely due to inequitable vaccine supply, weak health systems, and vaccine hesitancy [4]. Sub-Saharan Africa lags behind the global average, with only about 36% of the population receiving at least one dose by 2023 [5]. Uganda rolled out its vaccination program in March 2021, with priority groups including health workers, teachers, and high-risk populations [6]. While first-dose coverage has been commendable, completion of the second dose remains sub-optimal, threatening the overall effectiveness of the program.

Despite strong government and partner-led efforts, completion rates for COVID-19 vaccination remain inadequate in Uganda. By early 2024, 82% of adults had received the first dose, but only 59% had completed the two-dose regimen [7]. In Masindi District, uptake of the first dose was notably high at 94%, yet second-dose coverage was only 47% [7], meaning nearly half of those who received the first dose failed to return for the second. This drop-off undermines herd immunity efforts, increases vulnerability to severe illness, and wastes limited vaccine resources. Limited evidence exists to explain why such a significant gap persists, especially in rural districts like Masindi, despite national campaigns, integration of COVID-19 vaccination into routine immunization, and ongoing risk communication efforts.

Several studies have identified multi-level determinants of COVID-19 vaccine uptake. At the individual level, factors such as risk perception, vaccine safety concerns, and misinformation contribute significantly to hesitancy [8, 9]. At the community level, cultural norms, privacy preferences, and trust in local leaders affect acceptance [10]. Health system factors, including vaccine availability, cold-chain logistics, and stockouts, also influence access and completion [11]. Studies in Uganda and neighboring countries show that while community-led mobilization and decentralization of services improve initial uptake, second-dose completion often lags due to reduced perceived risk, logistical barriers, and inadequate follow-up mechanisms [12–13]. However, limited research has specifically focused on second-dose uptake in rural Ugandan settings, creating a knowledge gap.

Understanding the barriers and facilitators to second-dose COVID-19 vaccine uptake in Masindi District is critical for optimizing vaccination strategies. High first-dose coverage demonstrates community willingness to participate, yet the sharp decline at the second dose suggests systemic, community, or behavioral barriers that need to be addressed. Identifying these factors can inform targeted interventions to improve second-dose and booster uptake, strengthen routine immunization integration, and enhance preparedness for future pandemics. The findings from this study will be useful for policymakers, district health authorities, and implementing partners in tailoring strategies that respond to local realities while aligning with national and global immunization goals.

## Materials and Methods

### Ethical considerations

This study was reviewed and approved by the Uganda Martyrs University Faculty of Health Sciences and the Mildmay Uganda research and Ethics Committee under reference number; MUREC-2024-521. Permission to conduct the study was also obtained from Masindi District Health Officer (DHO) and the District Health Team (DHT). The study was conducted in accordance with the ethical principles of the Declaration of Helsinki. Before recruitment, the purpose and procedures of the study were explained to potential participants, and participation was voluntary. Written informed consent was obtained from all participants prior to enrolment into the study. For participants who were unable to read or write, the consent form was read aloud in the local language, adequate time was provided for questions, and informed consent was documented using a thumbprint in the presence of an impartial witness who also signed the consent form. Confidentiality was maintained throughout the study, and all information collected was used solely for research purposes.

### Study design and setting

This was a community-based quantitative cross-sectional study conducted using a structured, pre-tested, interviewer-administered questionnaire to assess factors associated with second-dose COVID-19 vaccine uptake among adults in Masindi district, Uganda. The district is located in mid-western Uganda, covering 18 sub-counties. Data collection took place across 20 villages selected from randomly chosen sub-counties and parishes, representing both rural and semi-urban settings.

### Study population

The study targeted adult residents of Masindi district aged 18 years and above who had received at least one dose of a COVID-19 vaccine, as this age group was the primary focus of national vaccination efforts due to higher COVID-19 morbidity and mortality [6].

### Inclusion criteria

The study included adult residents of Masindi district aged 18 years and above who had received at least one dose of a COVID-19 vaccine, resided in the selected households for at least 12 months and provided consent.

### Exclusion criteria

The study excluded individuals who were severely ill or cognitively unable to respond to the questionnaire.

### Sample size determination

The sample size formula for Kish Leslie (1965) was used as expressed below, Sample size,

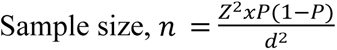

Where Z is the standard normal deviate of 1.96 (95% confidence interval)

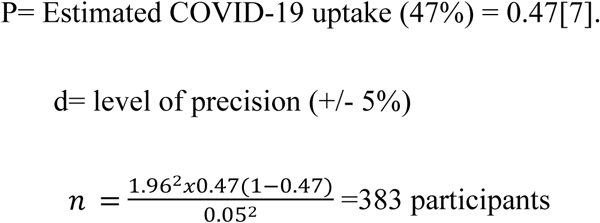

Considering a non-response rate of 10%

Final sample size= Effective sample size/ (1-non response rate anticipated)

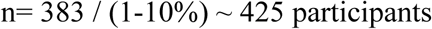

To account for cluster sampling, a design effect of 1.5 was applied, consistent with community-based surveys where intra-cluster correlation is expected.

Therefore, considering a design effect of 1.5, n=637

### Sampling procedure

Multi-stage random sampling was used to select study respondents. Masindi district is divided into 18 sub-counties. Five of the 18 sub-counties were selected by simple random sampling technique using lottery method. This was done by writing all the names of the sub-counties on individual pieces of paper, folded, mixed and placed in a container, then five sub-counties drawn randomly without replacement. This sampling technique was also applied at sub-county level and parish level to select two parish from each sub-county and two villages from each parish to participate in the study. From each of the selected sub-counties, two parishes were selected by simple random sampling. From each village, a list of households was obtained and systematic random sampling was used to select households. In households with more than one eligible adult, simple random selection was applied to select one respondent. The number of participants selected per village was determined using probability proportional to size (PPS) based on the estimated number of adults per village. Village population data were obtained from the Masindi district population officer (Table 1).

**Table 1:** Sampling criteria for study participants.

| Village | Sampling frame (adults per village) | Proportion of eligible participants | Sample size |
| --- | --- | --- | --- |
| Bwinamira | 1174 | 0.0684 | 44 |
| Lugazi | 774 | 0.0451 | 29 |
| Bulyango | 532 | 0.0310 | 20 |
| Waipacu | 746 | 0.0435 | 28 |
| Maiha | 771 | 0.0449 | 29 |
| Musoma | 427 | 0.0249 | 16 |
| Biseke | 600 | 0.0350 | 22 |
| Kyabakooto | 500 | 0.0291 | 19 |
| Kidamuke | 1408 | 0.0821 | 52 |
| Kapeeka1 | 674 | 0.0393 | 25 |
| Ewafala | 860 | 0.0501 | 32 |
| Bisenyi | 2090 | 0.1218 | 78 |
| Kigezi | 999 | 0.0582 | 37 |
| Kizinga | 796 | 0.0464 | 30 |
| Kayera | 1026 | 0.0598 | 38 |
| Matuugo | 566 | 0.0330 | 21 |
| Kiyuya | 983 | 0.0573 | 36 |
| Kyamudikya | 845 | 0.0493 | 31 |
| Kasomoro | 732 | 0.0427 | 27 |
| Katumba | 653 | 0.0381 | 24 |
| **Total** | **17156** | **1.000** | **637** |

*(Data from Masindi district population office, 2025).*

### Study variables

The primary outcome variable was COVID-19 second dose vaccine uptake, measured by whether individuals had received the second dose. Respondents were asked directly if they had completed the second dose, with responses categorized as “Yes” or “No,” reflecting completion or non-completion of the vaccine series. Independent variables included socio-demographic (age, gender, education level, occupation, marital status, and income), individual (perceived safety and efficacy of the vaccine, attitudes toward vaccination, and perception of disease severity), health system (distance to vaccination sites, vaccination related costs, vaccine availability), and community-level factors (political and media influence, cultural beliefs, peer influence, residence (urban/rural)).

### Data collection

Trained research assistants with support from the principal investigator collected data for a period of one month from 10 April 2025 to 10 May 2025 using a pre-tested interviewer administered structured questionnaire. Local leaders and village health teams only facilitated community entry and did not participate in data collection to minimize social desirability bias. The questionnaire was adapted from validated tools used in a similar study [14] and customized to meet the study objectives. The questionnaire covered socio-demographic characteristics, COVID-19 vaccine second dose uptake, individual factors, health system factors, and community-related factors

### Training of research assistants

Prior to data collection, the principal investigator selected the research assistants, trained them for four days on questionnaire administration, data collection, and ethical considerations; they were also trained in effective communication skills, obtaining informed consent, maintaining confidentiality, and addressing participants’ questions about COVID-19 vaccination in a neutral, respectful, and non-judgmental manner. The research assistants were aged between 20 and 30 years and fluent in the local language and attained at least certificate qualification with some holding diploma and bachelor’s degree in health-related or social science field. This helped to ensure free, open communication for the participants and reliable administration of the questionnaire.

### Quality control measures

The questionnaire was translated into Runyoro and subsequently back-translated into English to verify semantic consistency. Pretesting was conducted among 25 adults with the history of having received at least one dose of COVID-19 vaccine in Hoima district, which has similar socio-demographic characteristics to Masindi. Internal consistency reliability was assessed using Cronbach’s alpha, yielding a coefficient of ≥0.86 across key domains, indicating good reliability. Feedback from pretesting informed refinement of question wording and sequencing. Fieldwork was overseen by the principal investigator to ensure completeness and accuracy of data collected.

### Data analysis

Data analysis was performed using Statistical Package for Social Sciences (SPSS) version 26. Descriptive statistics were used to summarize participants’ characteristics, with categorical variables presented as frequencies and percentages. Prior to multivariable modeling, multicollinearity among independent variables was assessed using the variance inflation factor (VIF), and all variables had VIF values below 5, indicating absence of significant multicollinearity. Bivariate analysis was conducted using modified Poisson regression to determine factors associated with COVID-19 vaccine second dose uptake. Variables with p-values less than 0.2 at bivariate analysis were considered for multivariable analysis to avoid excluding potential predictors [15]. Modified Poisson regression with robust standard errors was used to estimate adjusted prevalence ratios (APRs) and 95% confidence intervals (CI) because the outcome was common (>10%). Model fit was assessed using the Omnibus test (X^2^ = 72.3, df = 36, p< 0.001), indicating a good model fit. Adjusted prevalence ratios (APRs) with 95% CI were reported, and statistical significance was considered at p<0.05.

## Results

### Sociodemographic characteristics of the respondents

A total of 637 respondents’ data were analyzed in this study. The mean age was 40.3 ± 15.4 years, and the median age was 40 years with an interquartile range (IQR) of 28-50 years. Most of the respondents (61.7%) resided in rural areas, and the majority (55.5%) were married. Regarding education, 27.9% had attained tertiary education, 26.1% primary education, 25.4% secondary education, and 20.4% had no formal education. About 61.2% of the participants were of low/poor economic status, and over 50.5% had 3 to 5 dependents. Majority of the participants were married (55.6%). The distribution across sub-counties was fairly balanced, with Budongo (23.1%), Miirya (22.3%), Kabango (21.0%), Kimengo (17.1%), and Bwijanga (16.5%). With regard to religion, 31.7% were Catholic, 28.1% Anglican, 25.3% Pentecostal, and 14.9% Muslim (Table 2).

**Table 2:** Socio-demographic characteristics of the respondents (N = 637)

| Variables | Frequency (n) | Percentage (%) |
| --- | --- | --- |
| **Age (Years)** |  |  |
| Age (Mean ± SD) | 40.3±15.4 | - |
| **Age (Median, IQR)** | 40 (28, 50) | - |
| ≤37 | 271 | 42.5 |
| 38-57 | 241 | 37.8 |
| ≥58 | 125 | 19.6 |
| **Marital status** |  |  |
| Not married | 187 | 29.4 |
| Married | 354 | 55.6 |
| Divorced | 70 | 11.0 |
| Widowed | 26 | 4.1 |
| **Sub-County** |  |  |
| Budongo | 147 | 23.1 |
| Bwijanga | 105 | 16.5 |
| Kabango | 134 | 21.0 |
| Kimengo | 109 | 17.1 |
| Miirya | 142 | 22.3 |
| **Religion** |  |  |
| Anglican | 179 | 28.1 |
| Catholic | 202 | 31.7 |
| Muslim | 95 | 14.9 |
| Pentecostal | 161 | 25.3 |
| **Level of education** |  |  |
| No formal education | 130 | 20.4 |
| Primary | 166 | 26.1 |
| Secondary | 163 | 25.6 |
| Tertiary | 178 | 27.9 |
| **Area of residence** |  |  |
| Rural | 379 | 59.5 |
| Urban | 258 | 40.5 |
| **Economic status** |  |  |
| Low | 390 | 61.2 |
| High | 247 | 38.8 |
| **No. of dependents** |  |  |
| ≤2 | 153 | 24.0 |
| 3-5 | 322 | 50.5 |
| ≥6 | 162 | 25.5 |

### Level of COVID-19 vaccine second dose uptake in Masindi district

Of the 637 adults who had received at least one dose of a COVID-19 vaccine, 280 (44.0%) had received the second dose

### Factors associated with COVID-19 vaccine second dose uptake in Masindi district (Unadjusted analysis)

#### Sociodemographic characteristics of study participants

Table 3 presents results of the bivariate analysis of socio-demographic factors associated with uptake of the second dose of the COVID-19 vaccine. Area of residence, and economic status were significantly associated with second dose uptake. Respondents residing in urban areas were significantly more likely to take the second dose compared to those in rural areas (CPR 2.1, 95% CI: 1.76-2.81, p < 0.001) with 64.0% of urban residents completing the second dose compared to 30.3% of rural residents. Participants with high economic status were 3.0 times more likely to have received the second dose compared to those with low economic status (CPR 3.0, 95% CI: 2.47-3.59, p < 0.001) (Table 3)

**Table 3.**
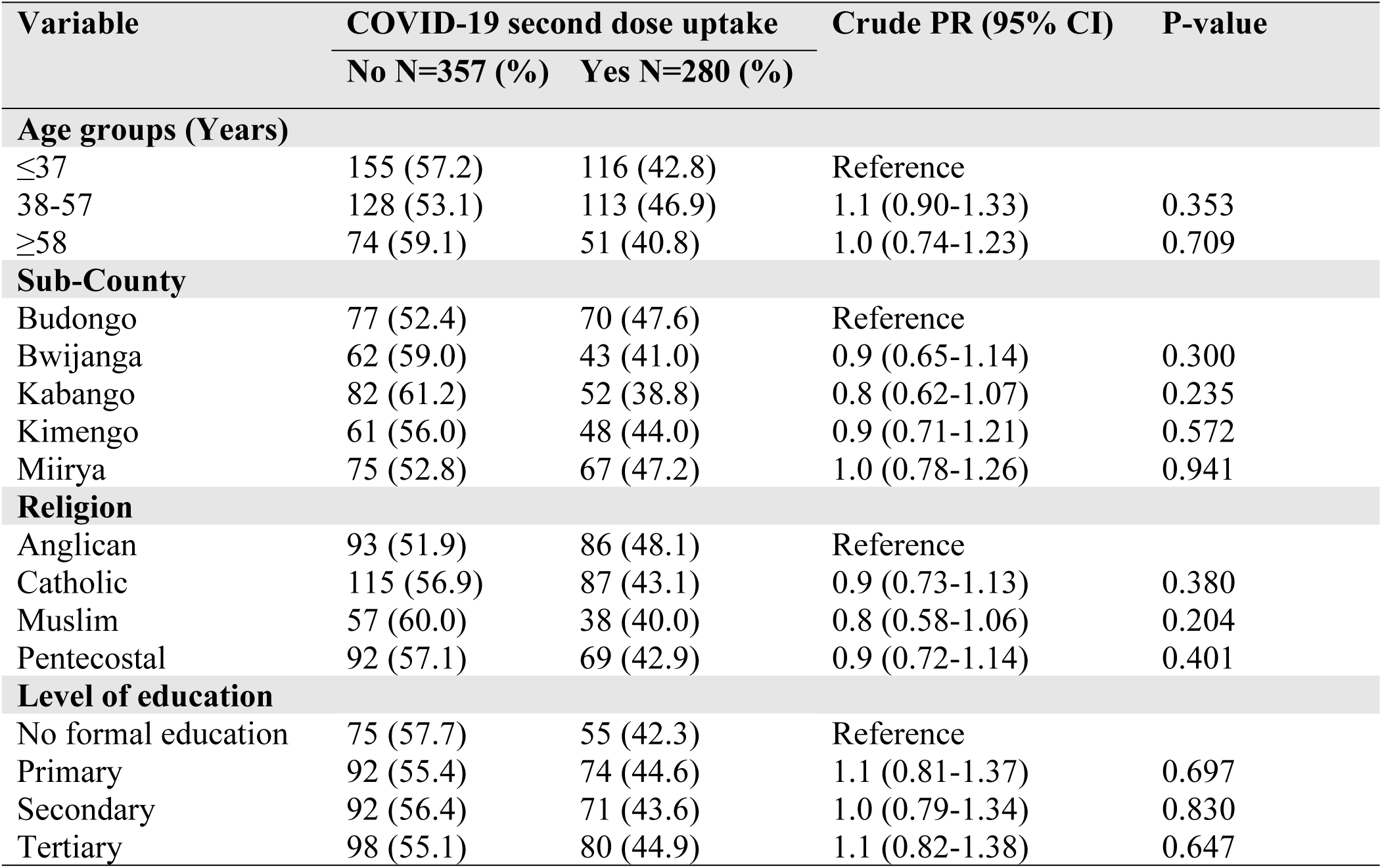
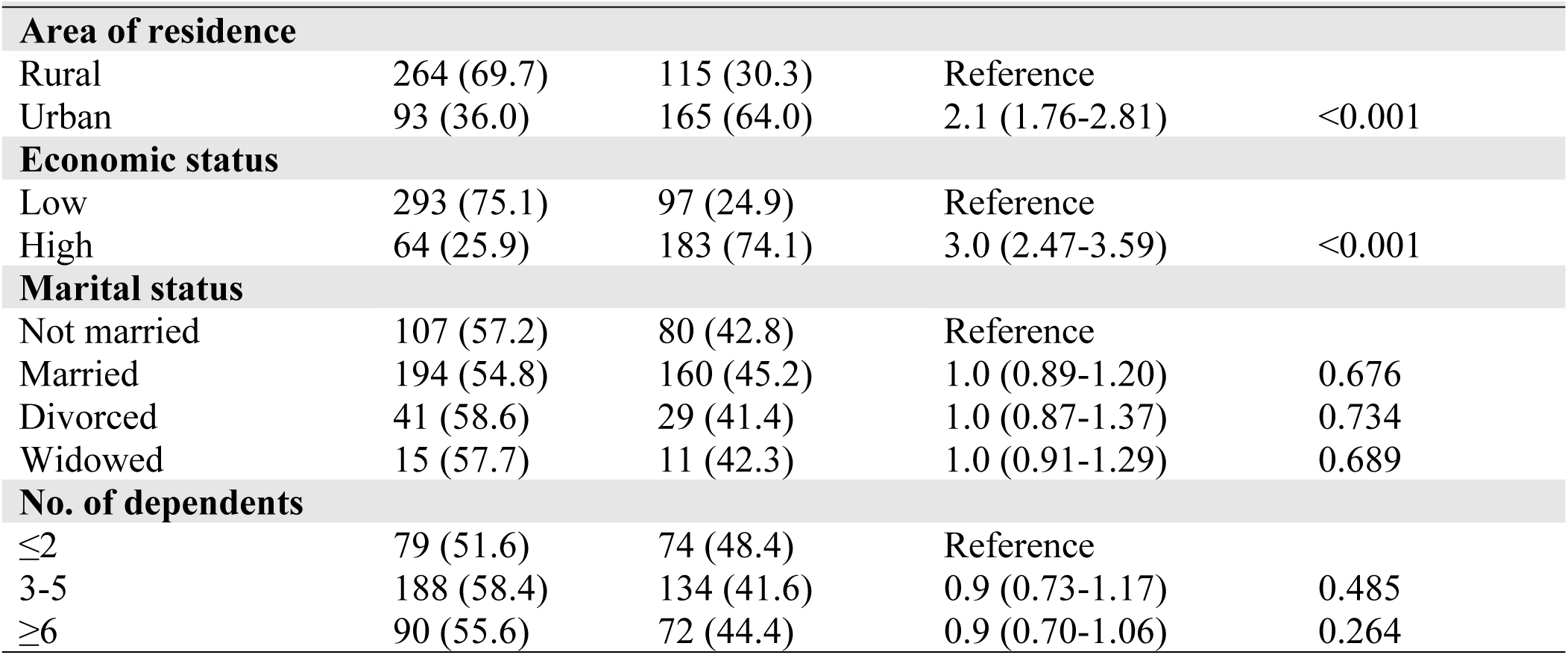
Unadjusted analysis of the respondent characteristics associated with COVID-19 second dose vaccine uptake.

### Individual factors

Several individual-level factors were significantly associated with uptake of the second dose of the COVID-19 vaccine. Participants who perceived themselves to be at high risk of contracting COVID-19 were 2.8 times more likely to receive the second dose compared to those who perceived a low risk (CPR 2.8, 95% CI: 1.96-3.64, p < 0.001). Experiencing adverse side effects after the first dose was negatively associated with uptake. Only 33.3% of respondents who experienced adverse effects received the second dose, compared to 73.1% among those who did not. Those who reported side effects were 30% less likely to receive the second dose (CPR 0.5, 95% CI: 0.39-0.63, p < 0.001).In addition, respondents with chronic illnesses were significantly more likely to receive the second dose (CPR 2.7; 95% CI: 2.00-3.37, p<0.001). (Table 4)

**Table 4.**
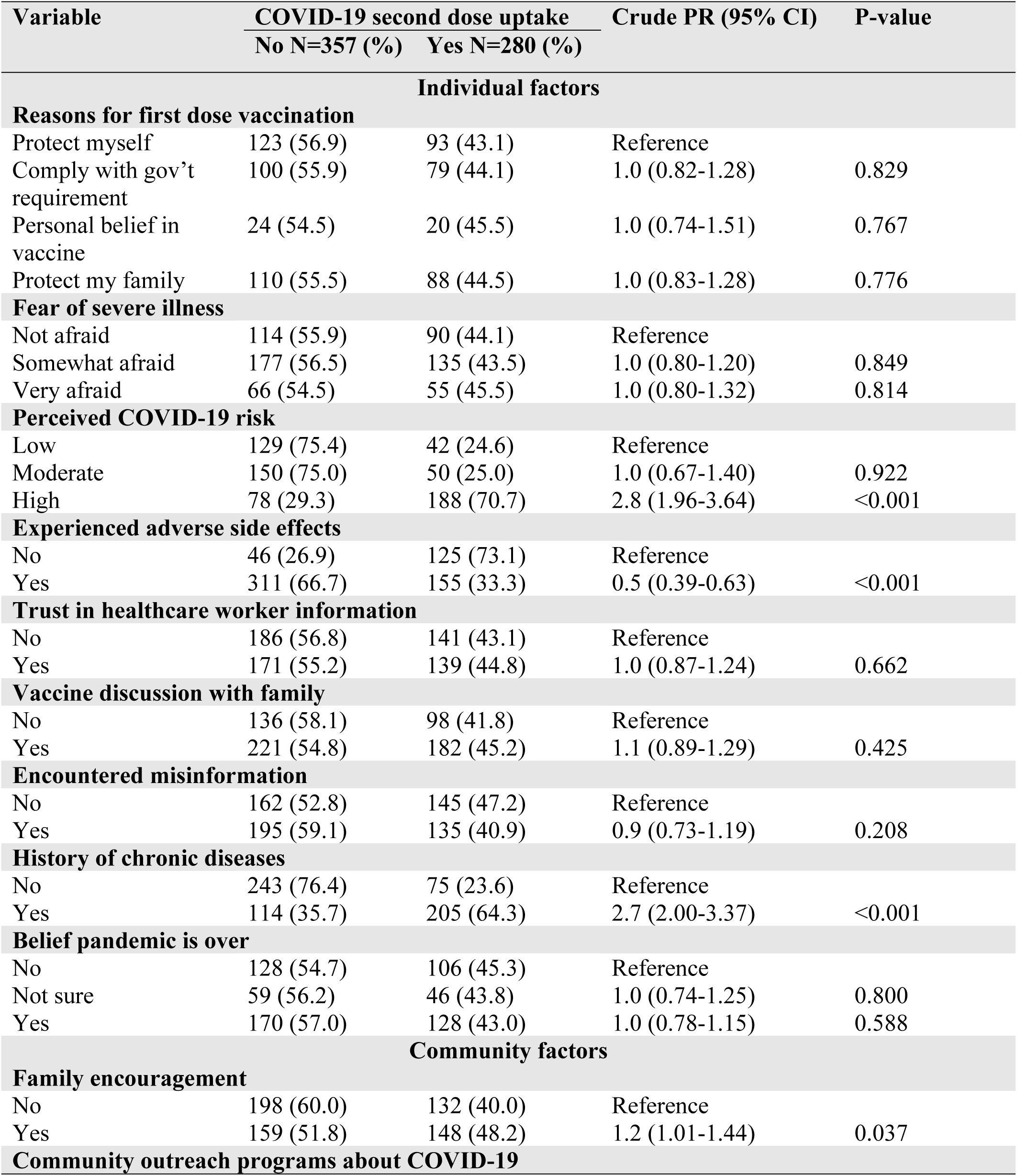
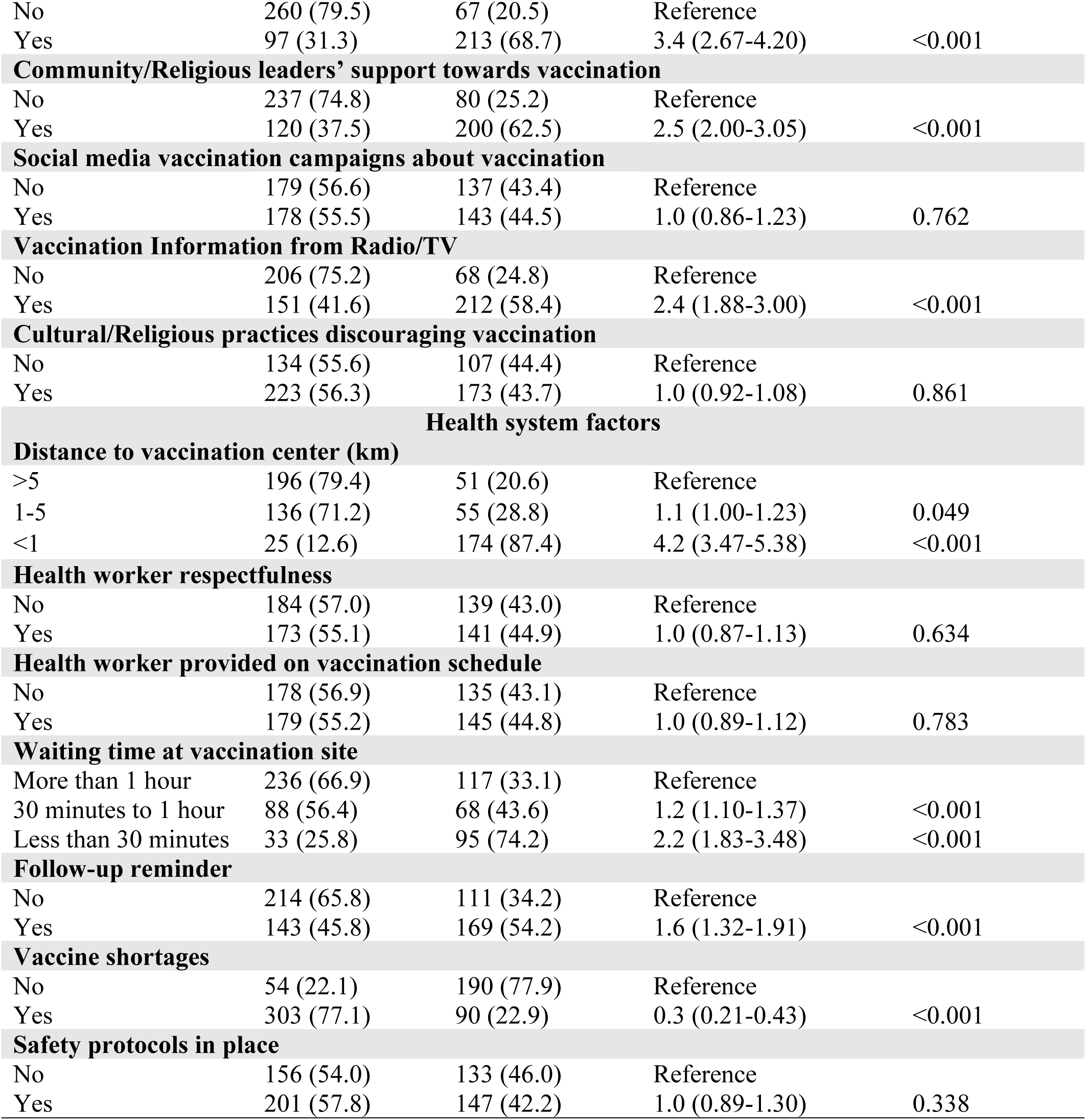
Unadjusted analysis of factors (Individual, Community and Health system) associated with COVID-19 second dose uptake (n=637)

### Community-level factors

Several community-level factors were significantly associated with second dose COVID-19 vaccine uptake. Respondents living in communities with COVID-19 outreach programs had substantially higher uptake (68.7%) compared to those in communities without such programs (20.5%). Those exposed to community outreach efforts were 3.4 times more likely to receive the second dose (CPR 3.4, 95% CI: 2.67-4.20, p < 0.001). Support from community and religious leaders was also positively associated with second dose uptake. Participants who reported support from these leaders were 2.5 times more likely to receive the second dose compared to those who did not (CPR 2.5, 95% CI: 2.00-3.05, p < 0.001). Those who were supported and encouraged by their family members to take the second dose were 1.2 times more likely to do as opposed to their counterparts who did not have the family support in form of encouragement (CPR 1.2, 95% CI: 1.01-1.44, p = 0.037). Exposure to vaccination information through radio or television significantly increased second dose uptake. Respondents who received such media information were 2.4 times more likely to receive a second dose compared to those who were not exposed (CPR 2.4, 95% CI: 1.88-3.00, p < 0.001). (Table 4)

### Health system–related factors

Several health system–related factors were significantly associated with uptake of the second dose of the COVID-19 vaccine. Participants who lived less than 1 km from a vaccination center were 1.9 times more likely to take the second dose compared to those who lived more than 5 km away (CPR 1.9, 95% CI: 1.72-2.88, p < 0.001). Participants who experienced vaccine shortages at health facilities had significantly lower uptake of the second dose (22.9%) compared to 77.9% among those who did not experience shortages. Those who reported vaccine shortages were 70% less likely to receive the second dose (CPR 0.3, 95% CI: 0.21-0.43, p < 0.001). Shorter waiting times were also associated with higher uptake. Respondents who waited less than 30 minutes were 2.2 times more likely to take the second dose than those who waited more than an hour (CPR 2.2, 95% CI: 1.83-3.48, p < 0.001). Likewise, those who waited between 30 minutes and 1 hour were 1.2 times more likely to take the second dose (CPR 1.2, 95% CI: 1.10–1.37, p < 0.001). Participants who received follow-ups reminder messages were also significantly more likely to take the second dose (CPR 1.6, 95% CI: 1.32-1.91, p < 0.001). (Table 4)

### Multivariable analysis

All variables that had P values <0.2 at bivariate level of analysis were selected for the multivariable analysis. The final model depicts the best fit for this analysis. All variables that remained in the multivariable model are statistically significant and they are thus significantly associated with COVID-19 vaccine second dose uptake among adults Masindi district.

### Factors independently associated with COVID-19 vaccine second dose uptake

The factors that remained significantly associated with increased likelihood of COVID-19 vaccine second dose uptake at multivariable analysis level included: high economic status (APR = 2.2, 95% CI: 1.16-3.07, p < 0.001), urban residence (APR = 1.3, 95% CI: 1.13-1.62, p < 0.001), high perceived risk of COVID-19 (APR = 2.0, 95% CI: 1.59-2.55, p = 0.001), chronic illness (APR 1.9; 95% CI: 1.60-2.33, p <0.001), living less than 1 km from a vaccination site (APR = 2.0, 95% CI: 1.49-3.01, p < 0.001), Shorter waiting times; under 30 minutes (APR = 1.7, 95% CI: 1.41-2.08, p < 0.001), community/religious leaders’ support (APR = 1.7, 95% CI: 1.41-2.16, p < 0.001), Community outreach programs (APR = 2.5, 95% CI: 1.99-3.36, p < 0.001), and Exposure to vaccination information through radio or TV (APR = 1.5, 95% CI: 1.25–1.88, p<0.001). Vaccine shortages (APR = 0.4, 95% CI: 0.31-0.55, p < 0.001), and experiencing of adverse side effects after the first dose (APR = 0.8, 95% CI: 0.71-0.97, p = 0.020) were associated with reduced prevalence of receiving a second dose (Table 5).

**Table 5.**
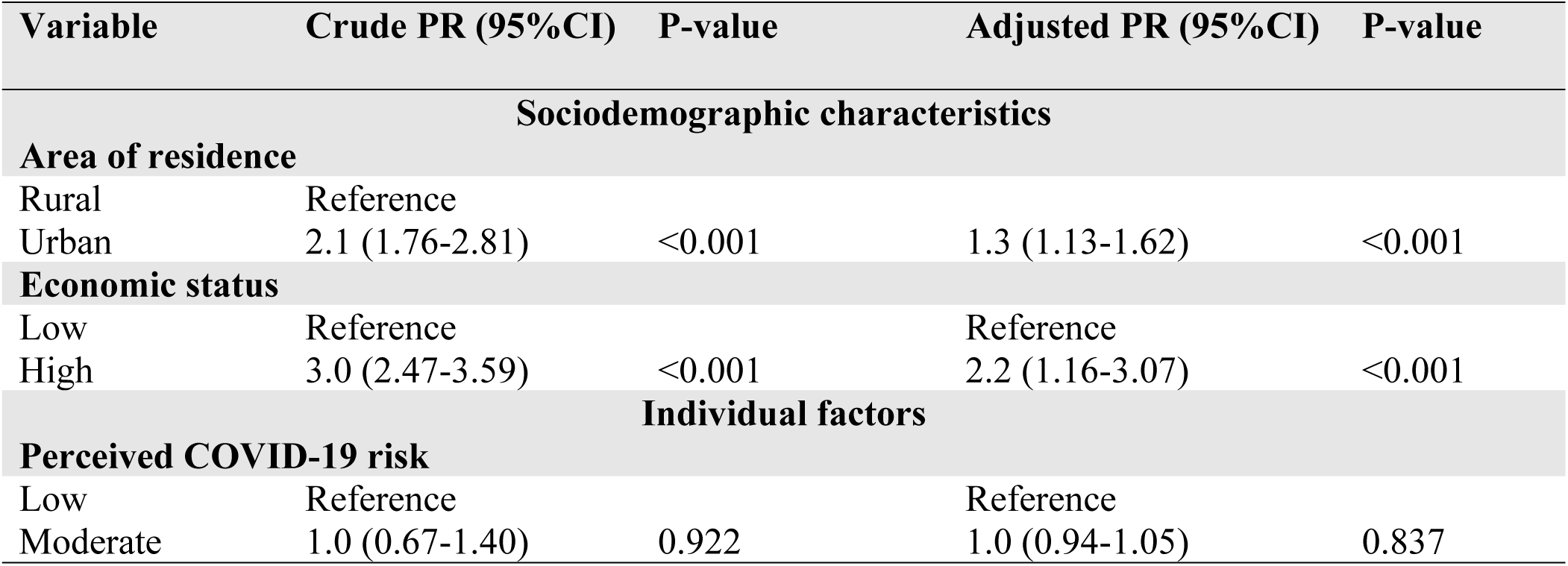
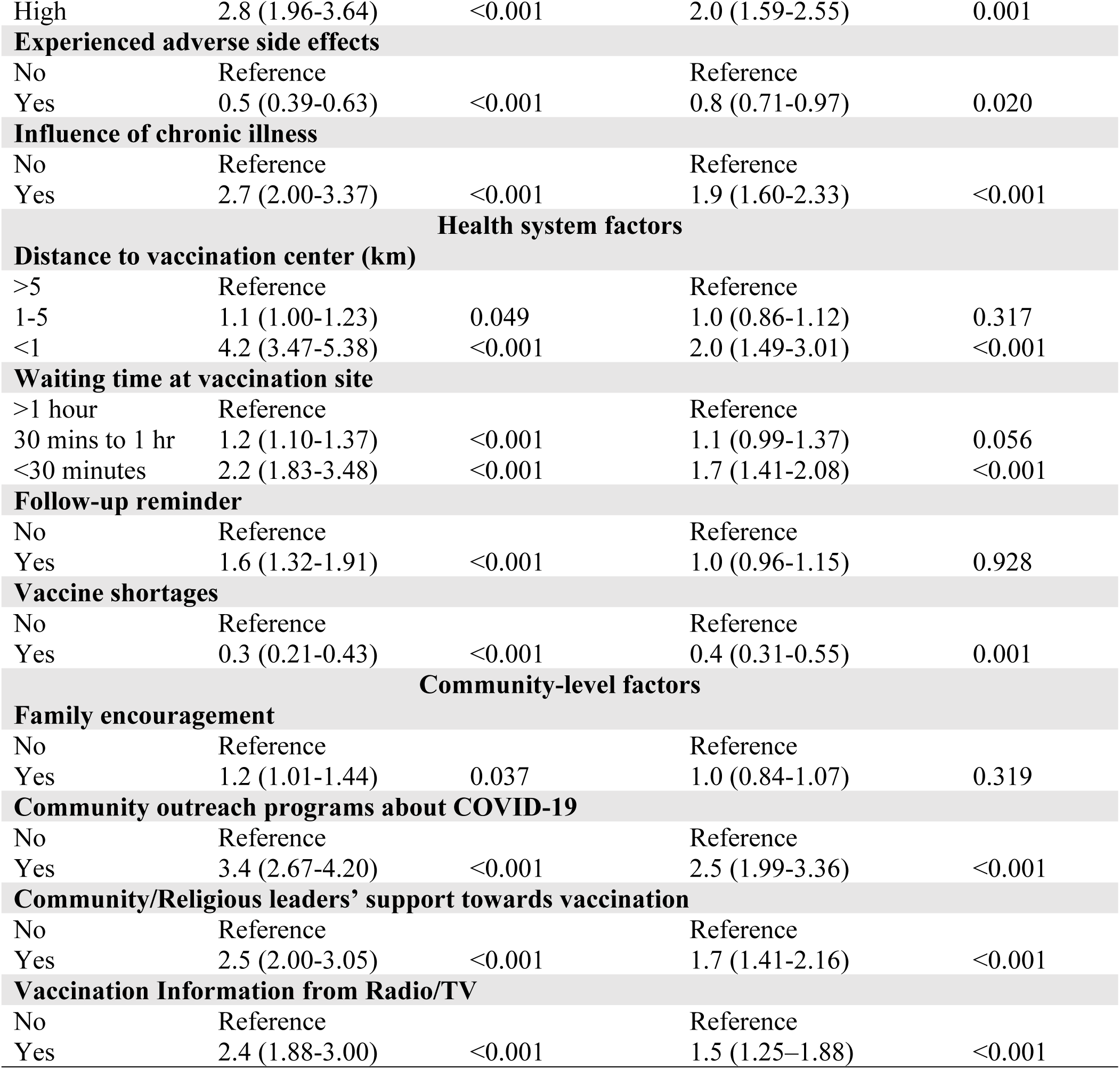
Multivariable analysis of factors associated with COVID-19 vaccine second dose uptake.

| Variable | Crude PR (95%CI) | P-value | Adjusted PR (95%CI) | P-value |
| --- | --- | --- | --- | --- |
| **Sociodemographic characteristics** |  |  |  |  |
| **Area of residence** |  |  |  |  |
| Rural | Reference |  |  |  |
| Urban | 2.1 (1.76-2.81) | <0.001 | 1.3 (1.13-1.62) | <0.001 |
| **Economic status** |  |  |  |  |
| Low | Reference |  | Reference |  |
| High | 3.0 (2.47-3.59) | <0.001 | 2.2 (1.16-3.07) | <0.001 |
| **Individual factors** |  |  |  |  |
| **Perceived COVID-19 risk** |  |  |  |  |
| Low | Reference |  | Reference |  |
| Moderate | 1.0 (0.67-1.40) | 0.922 | 1.0 (0.94-1.05) | 0.837 |
| High | 2.8 (1.96-3.64) | <0.001 | 2.0 (1.59-2.55) | 0.001 |
| **Experienced adverse side effects** |  |  |  |  |
| No | Reference |  | Reference |  |
| Yes | 0.5 (0.39-0.63) | <0.001 | 0.8 (0.71-0.97) | 0.020 |
| **Influence of chronic illness** |  |  |  |  |
| No | Reference |  | Reference |  |
| Yes | 2.7 (2.00-3.37) | <0.001 | 1.9 (1.60-2.33) | <0.001 |
| **Health system factors** |  |  |  |  |
| **Distance to vaccination center (km)** |  |  |  |  |
| >5 | Reference |  | Reference |  |
| 1-5 | 1.1 (1.00-1.23) | 0.049 | 1.0 (0.86-1.12) | 0.317 |
| <1 | 4.2 (3.47-5.38) | <0.001 | 2.0 (1.49-3.01) | <0.001 |
| **Waiting time at vaccination site** |  |  |  |  |
| >1 hour | Reference |  | Reference |  |
| 30 mins to 1 hr | 1.2 (1.10-1.37) | <0.001 | 1.1 (0.99-1.37) | 0.056 |
| <30 minutes | 2.2 (1.83-3.48) | <0.001 | 1.7 (1.41-2.08) | <0.001 |
| **Follow-up reminder** |  |  |  |  |
| No | Reference |  | Reference |  |
| Yes | 1.6 (1.32-1.91) | <0.001 | 1.0 (0.96-1.15) | 0.928 |
| **Vaccine shortages** |  |  |  |  |
| No | Reference |  | Reference |  |
| Yes | 0.3 (0.21-0.43) | <0.001 | 0.4 (0.31-0.55) | 0.001 |
| **Community-level factors** |  |  |  |  |
| **Family encouragement** |  |  |  |  |
| No | Reference |  | Reference |  |
| Yes | 1.2 (1.01-1.44) | 0.037 | 1.0 (0.84-1.07) | 0.319 |
| **Community outreach programs about COVID-19** |  |  |  |  |
| No | Reference |  | Reference |  |
| Yes | 3.4 (2.67-4.20) | <0.001 | 2.5 (1.99-3.36) | <0.001 |
| **Community/Religious leaders' support towards vaccination** |  |  |  |  |
| No | Reference |  | Reference |  |
| Yes | 2.5 (2.00-3.05) | <0.001 | 1.7 (1.41-2.16) | <0.001 |
| **Vaccination Information from Radio/TV** |  |  |  |  |
| No | Reference |  | Reference |  |
| Yes | 2.4 (1.88-3.00) | <0.001 | 1.5 (1.25-1.88) | <0.001 |

## Discussion

In this study, 44.0% of participants received their second dose of the COVID-19 vaccine. This indicates that less than half of the adult participants had completed their second dose. This observation suggests a potential discrepancy between starting COVID-19 vaccination and finishing the recommended primary vaccination schedule. A comparable trend was noted in a population-based study in Uganda’s slum and estate communities, with primary vaccination series completion rates of 43.8% and 39.9%, respectively [16]. While these results are generally similar, variations in study populations, contexts, and definitions of outcomes should be taken into account when interpreting these figures. The study’s results indicate that incomplete vaccination was observed beyond just Masindi District or a specific Ugandan community. Several factors may affect the completion of a vaccination series, such as the accessibility of vaccination services, vaccine supply, the expense of time and travel, and worries regarding side effects. This study’s findings, specifically the links between receiving the second vaccine dose and the distance to vaccination centers, wait times, vaccine availability, and reactions after the first dose, offer local evidence of potential obstacles to completing vaccination. These observations highlight the need to address both limitations in service provision and individual concerns.

People with higher incomes were more likely to get their second vaccine dose. This association suggests that the ability to meet the indirect costs of vaccination may influence completion, even where the vaccine itself is provided without charge. Indirect expenses such as transport cost, time away from income-generating activities and competing household responsibilities may impose a greater burden on individuals with fewer resources especially when the vaccination requires a return visit. Similar trends have been noted both in United States, Nigeria and South African where individuals with higher economic status were most likely to complete a COVID-19 vaccination series [17, 18]. Taken together, this evidence supports the need to address the practical costs of accessing vaccination rather than relying exclusively on information campaigns.

Geographical accessibility may influence vaccination completion by affecting the time, transport costs and effort required to return for their follow-up dose. Evidence from six sub-Saharan African countries indicates that long distances to vaccination points can prevent people who are willing to be vaccinated from accessing services [19]. Similarly, a scoping review of COVID-19 vaccination research in sub-Saharan Africa identified travel distance as a barrier to vaccine dose completion [20]. These findings concur with the assertion of WHO that convenience of logistics such as proximity may influence an individual’s ability to complete the vaccination series as distant locations and extended queues may lead opportunity costs, especially to individuals with daily income-generating pursuits [21]. To increase second dose uptake, health planners should prioritize user-centered service design and should also invest in mobile vaccination teams that extend coverage to under-served populations. Evidence from a cluster-randomised trial in Sierra Leone further demonstrates that bringing vaccination teams and doses directly to remote communities can increase vaccine uptake [22].

Urban residents may be more likely to complete the COVID-19 immunization schedule due to their closer proximity to medical facilities, more frequent interactions with healthcare professionals, and easier access to vaccination information. However, obstacles like work obligations and a lack of time to attend immunization appointments may also affect urban residents. Research from six sub-Saharan African nations, including Uganda, shows that vaccination obstacles differ between rural and urban environments, with work-related limitations more prevalent in urban areas and distance and vaccine availability more prominent in rural regions [19]. These findings emphasize the significance of customizing immunization programs to meet local needs, such as enhancing outreach in rural areas and offering urban inhabitants easy immunization chances.

Perceived risk may influence vaccination decisions by shaping how seriously individuals regard the consequences of infection and how much benefit they expect from completing the recommended schedule. This interpretation is consistent with the Health Belief Model, which proposes that perceived susceptibility and severity can influence preventive health behaviour [23]. Similar relationships between perceived risk and vaccination behaviour have been reported in study from Taiwan [24]. The systematic review on behavioural responses during the pandemic, identified risk perception as one of the psychological factors relevant to protective behavior [25]. However, risk perception alone may not be sufficient to support vaccination completion. Even individuals who recognize the potential consequences of COVID-19 may face practical barriers, have concerns about vaccine safety or encounter difficulties accessing subsequent doses. This highlights the need to address both perceptions of disease risk and the practical circumstances in which vaccination decisions are made.

The positive association between chronic illness and second-dose uptake may reflect greater perceived vulnerability to severe COVID-19 or more frequent contact with healthcare services among people managing long-term conditions. People living with chronic conditions interact more with healthcare workers while managing their conditions hence the increased opportunities of receiving about potential consequences of COVID-19. Our findings are in agreement with results from similar studies in Ethiopia and Australia about serious underlying medical conditions and COVID-19 second dose vaccine hesitancy [26, 27]. These findings highlight the potential role of perceived susceptibility to severe disease and concerns about the consequences of infection in shaping vaccination-related decisions among individuals with chronic conditions. Taken together, the evidence suggests that routine healthcare encounters may provide important opportunities to reinforce the benefits of completing the recommended vaccination schedule, particularly among people living with chronic illnesses.

The association between shorter waiting times and second-dose uptake in Masindi suggests that the efficiency of vaccination services may influence completion of the COVID-19 vaccination schedule. Long queues can impose opportunity costs on individuals who must balance healthcare visits with work, caregiving and other daily responsibilities, potentially discouraging them from seeking vaccination. A study in the United States of America (USA) about factors associated with delayed or missed second-dose cited shorter waiting time at the vaccination centers as being associated with increased chances of receiving the vaccine [28]. Similarly, a qualitative study among healthcare workers and older people in Uganda identified long queues at vaccination centers as a structural barrier to COVID-19 vaccine uptake [29]. Health facilities should adopt queue management systems, fast-track lines for follow-up doses, and consider appointment-based vaccination to improve flow. Training more vaccinators or reallocating staff during peak hours can also reduce congestion. Taken together, these findings suggest that reducing waiting times and improving the efficiency of vaccination services may help minimise practical barriers to vaccine access.

The negative association between experiencing adverse effects after the first dose and second-dose uptake suggests that post-vaccination experiences may influence decisions about completing the vaccination schedule. Concerns about vaccine safety and anticipated side effects have been identified as factors associated with COVID-19 vaccine hesitancy in a systematic review of vaccine uptake determinants [30]. Similarly, a qualitative study in southwestern Uganda found that reported post-vaccination symptoms and concerns about perceived adverse events influenced some participants’ willingness to receive additional doses [31]. These findings highlight the importance of providing clear counselling on expected post-vaccination symptoms, explaining when clinical assessment is necessary, and giving individuals the opportunities to discuss their concerns with qualified health workers. Acknowledging reported experiences while providing accurate, evidence-based safety information may help address concerns and support completion of the recommended vaccination schedule.

The negative association between vaccine shortages and the uptake of second doses underscores the necessity of ensuring consistent vaccine availability to complete vaccination schedules. Instances of stock-outs may lead to multiple visits, elevate transportation expenses, and diminish trust in vaccination services. A qualitative study conducted in Uganda recorded instances of missed vaccinations and the necessity for return visits due to the unavailability of vaccines, while regional data pointed to challenges in vaccine supply and distribution during the COVID-19 vaccination campaign [29, 32]. Additionally, research on routine immunization in Uganda emphasizes the critical nature of efficient vaccine distribution and inventory management [33]. In Masindi district, enhancing stock oversight, demand forecasting, and distribution, as well as effectively communicating vaccine availability, could help minimize missed opportunities.

Community outreach initiatives and second-dose COVID-19 vaccine uptake are positively correlated, indicating that bringing vaccination services and information closer to communities may help remove obstacles to finishing the immunization schedule. Outreach can lessen travel to permanent medical facilities, increase awareness, and address safety concerns. Our findings are in agreement with findings from the previous studies that documented that the use of mobile and outreach services to improve vaccine delivery [29, 34]. The results indicate that outreach efforts could be especially beneficial in areas where geographic barriers and the arrangement of regular services hinder vaccination. In Masindi District, enhancing outreach via community health workers, local leaders, and current health facility initiatives could create chances to inform individuals about the availability of subsequent doses and assist adults who struggle to access static vaccination locations.

Community and religious leaders may influence vaccination decisions because they can communicate health messages through trusted social networks and help address concerns about vaccination. Our finding suggest that social norms, trust, and media of communication play the pivotal role in shaping health behavior amidst a pandemic. It makes sense in a context like Uganda, where community leaders are the respected gatekeepers of information and decision-making within communities. Their approval not only attests to the credibility of public health initiatives but also dispels mistrust and uncertainty by followers. This finding is consistent with Ugandan studies, where religious and community leader endorsement boosted vaccine uptake among hesitant populations [35]. Similarly, in Pakistan, vaccine uptake was greatly improved upon once local imams were enlisted in vaccine promotion initiatives [36].

Access to accurate vaccination information through mainstream media may improve the completion of the COVID-19 vaccination series. Radio may be the most accessible and trustworthy source of information in the majority of rural and peri-urban areas throughout Uganda. If well harnessed, it can smother myths, send reminders about deadlines, and popularize vaccine acceptance. This finding is consistent with studies in Zambia, whereby media exposure to health messages from the television and radio significantly increased the chances of full COVID-19 vaccination [37]. A study in several sub-Saharan African settings reported that the absence of a trusted source of COVID-19 vaccine information was associated with vaccine hesitancy among adults [38]. Use of interaction with local radio stations for telephone call-in programs and talk shows is also an opportunity for the community members to ask questions and clarify concerns, hence gaining trust and transparency. Media communication should therefore be considered alongside service accessibility, vaccine availability and interpersonal engagement with health workers and community leaders.

### Public health implications for Masindi district and Uganda

The findings from our study have implications for national immunization planning as well as district-level service delivery. First, the association between economic status, place of residence, and accessibility to immunization services indicates that groups with real-world obstacles to receiving care should be given priority while planning for vaccination campaigns. Monitoring at the district level could help pinpoint areas with poor completion rates and guide the placement of outreach programs. Secondly, the links between vaccine shortages and waiting times highlight the importance of community demand alongside the quality and consistency of services. Thus, ensuring a steady supply of vaccines, facilitating service delivery, and providing clear information about vaccination protocols should all be integrated into initiatives aimed at enhancing completion rates. Third, the associations with perceived risk, adverse effects in the past, chronic illness, and community communication highlight the need of customized communication. Healthcare providers ought to be ready to alleviate concerns regarding vaccine safety and the consequences of inadequate immunization. Local media, religious figures, and community leaders can assist in disseminating accurate information and directing individuals toward available services. Additionally, connecting vaccination options with current health service interactions, such as chronic disease management, may also facilitate access for those who could gain from further guidance and easier availability.

At the national level, these results reinforce the broader concept that vaccination oversight should differentiate between initial uptake and the completion of multi-dose schedules. Programs that document the administration of the first dose without effectively tracking subsequent doses may miss populations that remain partially vaccinated. Enhancing follow-up, monitoring the availability of services, and investigating variations in completion rates based on socioeconomic and geographical factors could assist Uganda in pinpointing areas where additional support is necessary. While this study focuses on Masindi District, the barriers and facilitators identified may guide the creation of locally tailored strategies in other contexts, as long as variations in population characteristics and health service organization are considered.

## Strengths and Limitations

The major strength of this study lies in its community-based cross-sectional design, which ensured a representative sample and provided insights into the real-world experiences of vaccine uptake among adults in Masindi District. The use of multi-stage random sampling may have enhanced the reliability of findings by capturing diverse population groups. Additionally, the study holistically examined determinants across individual, health system, and community levels, allowing for a comprehensive understanding of barriers and facilitators of second dose uptake. Despite its strengths, the study had several limitations. The cross-sectional design limited the ability to establish causal relationships between the identified predictors and vaccine uptake. Reliance on self-reported data introduced potential recall and social desirability biases, as some respondents may have overreported vaccination status or provided responses they deemed socially acceptable. Furthermore, the study was confined to Masindi district, which may limit the generalizability of findings to other regions with different socio-economic or health system contexts. A longitudinal or mixed-methods design, including qualitative inquiry, would provide deeper insights into the evolving barriers to vaccine completion.

## Conclusion and recommendations

This study revealed that the uptake of the second dose of the COVID-19 vaccine in Masindi District was 44%, indicating that more than half of the study participants had not received the COVID-19 second dose. Second dose uptake was independently associated with socioeconomic status, perceived risk of COVID-19, chronic illness, geographical accessibility, waiting times, vaccine availability, adverse effects following the first dose, community outreach, support from community and religious leaders, and exposure to radio and television messages.

These findings emphasize that improving vaccine completion requires an integrated, multi-level approach addressing socioeconomic, individual, community and health-system factors. Henceforth, both district and national stakeholders should strengthen community outreach, ensure reliable vaccine supplies, reduce waiting times and geographical barriers, and provide clear information about vaccine safety and the importance of completing subsequent doses. These findings may inform locally tailored strategies to improve multi-dose vaccine completion in Masindi district and other settings with similar challenges.

## Data Availability

All raw data required to reproduce the results of this study are included within the Supporting Information files (S1 File).

## List of abbreviations and acronyms

CPR: Crude Prevalence Ratio
APR: Adjusted Prevalence Ratio
COVID-19: Corona Virus Disease 2019
IQR: Interquartile Range
SD: Standard Deviation

## Declarations

## Consent for publication

Not applicable.

## Author contributions

R.K and A.A conceived the study, designed the research, collected the data, and drafted the manuscript. R.K, L.N.Y.K, S.G.S, A.A and, R.K supervised, provided critical revisions, and provided expert guidance throughout the study design, data interpretation, and manuscript preparation. R.K, A.A, and S.G.S analyzed the data, developed the visualizations, and wrote the results section of the manuscript. All authors read and approved the final manuscript for submission.

## Acknowledgment.

We acknowledge the Masindi District administrative authorities and the District Health Team for granting us permission to conduct the study.

## Supporting Information

**S1 File. Dataset used for analysis.**

An anonymized dataset containing variables used in the quantitative analysis of the level of uptake and associated factors of COVID-19 second dose vaccine uptake in Masindi.

